# CanBART improves cross-platform prediction with a foundation model of targeted genomic panels

**DOI:** 10.1101/2025.11.04.25339512

**Authors:** Dmitrii Chebanov, Quaid Morris

## Abstract

Real-world genomic cohorts remain limited in size, unevenly profiled, and especially sparse for rare cancers. Many sequencing panels differ in gene coverage, leaving key molecular features unobserved and complicating cohort comparison, biomarker discovery, and clinical-trial enrollment.

CanBART, a generative foundation model trained on 144,000 tumor profiles, learns tokenized representations of somatic alterations and supports genomic completion, tumor classification, and synthetic-patient generation via a masked-language architecture.

We simulated a panel reduction scenario by limiting each MSK-IMPACT profile to the smaller set of genes measured in the DFCI OncoPanel, and asked the model to reconstruct the missing genes. In this setting, CanBART recovered over one-third of the held-out alterations with high confidence. Using sampling strategies adapted from NLP, we also generated biologically coherent “plausible patients” that expanded rare cancer cohorts, and training classifiers on these synthetic profiles improved accuracy for two-thirds of tumor types, particularly those with only 20-500 samples.

By completing missing genomic features and generating “plausible” synthetic patients, CanBART provides a scalable tool for rare cancer research and cross-panel harmonization.

## Introduction

Targeted panel sequencing has become a routine component of clinical oncology practice, informing treatment selection through actionable biomarkers and genomically-targeted therapies. In contrast to whole-genome or whole-exome sequencing, panel-based assays are optimized for speed and cost, and are therefore widely adopted in real-world settings. However, substantial heterogeneity exists across panels: different institutions profile different gene sets, panel sizes vary, and coverage evolves over time. Although these panels largely overlap in their core cancer genes, their non-identical design introduces systematic missingness and fragmentation in patient-level genomic profiles, complicating cross-institution dataset integration.

At the same time, the widespread clinical use of panel sequencing has produced large, richly annotated real-world datasets, motivating the development of computational approaches that can predict clinically actionable features and reveal molecular insights into cancer biology. Beyond treatment selection, one of the established applications of genomic profiling is tumor classification, particularly in challenging cases such as cancers of unknown primary (CUP), where conventional diagnostic evaluations can remain indeterminate. In these settings, even imperfect genomic classification can be clinically valuable: it can narrow plausible diagnoses, support confirmation of suspected origins, and guide therapy choices when histology and imaging are inconclusive. In practice, heterogeneity across real-world sequencing panels limits the reliability and portability of existing models. Tools capable of both inferring missing alterations and classifying tumors from partial profiles would therefore be highly valuable in clinical practice, enabling cross-platform prediction and more consistent use of real-world genomic cohorts.

Recent advances in large pre-trained models suggest that biological sequence data can support powerful representation-learning approaches. Models such as AlphaFold for protein structure prediction, Enformer for regulatory sequence modeling, and RNA-FM for transcriptome-level embeddings demonstrate that foundation model approaches can extract biologically meaningful structure directly from raw sequence-like inputs (1, 2, 3, 4).

A central motivation for this study was to develop a foundation model in which the tokens correspond to molecular alterations found in individual cancer patients. Inspired by natural language processing, we hypothesized that a generative Transformer architecture could be used not only for classification but also for creating realistic synthetic patient cohorts, helping to overcome the scarcity of labeled data for rare cancers. A further motivating precedent comes from the GDD-ENS study by Darmofal et al., which showed that classifiers trained exclusively on common cancers could predict the organ system of origin for previously unseen rare cancers with meaningful accuracy (5), suggesting shared latent genomic patterns between common and rare tumor types.

To explore this further, we introduce CanBART, a foundation model trained using the BART architecture, where the input “sentences” consist of molecular alterations identified in tumor profiling (Fig. 1A). The model is pre-trained using masked-language modeling on more than 144,000 cancer patient profiles and is subsequently fine-tuned for tumor classification, used to assess the biological meaningfulness of patient-level genomic representations and their utility for generation-based augmentation.

**Fig. 1.**
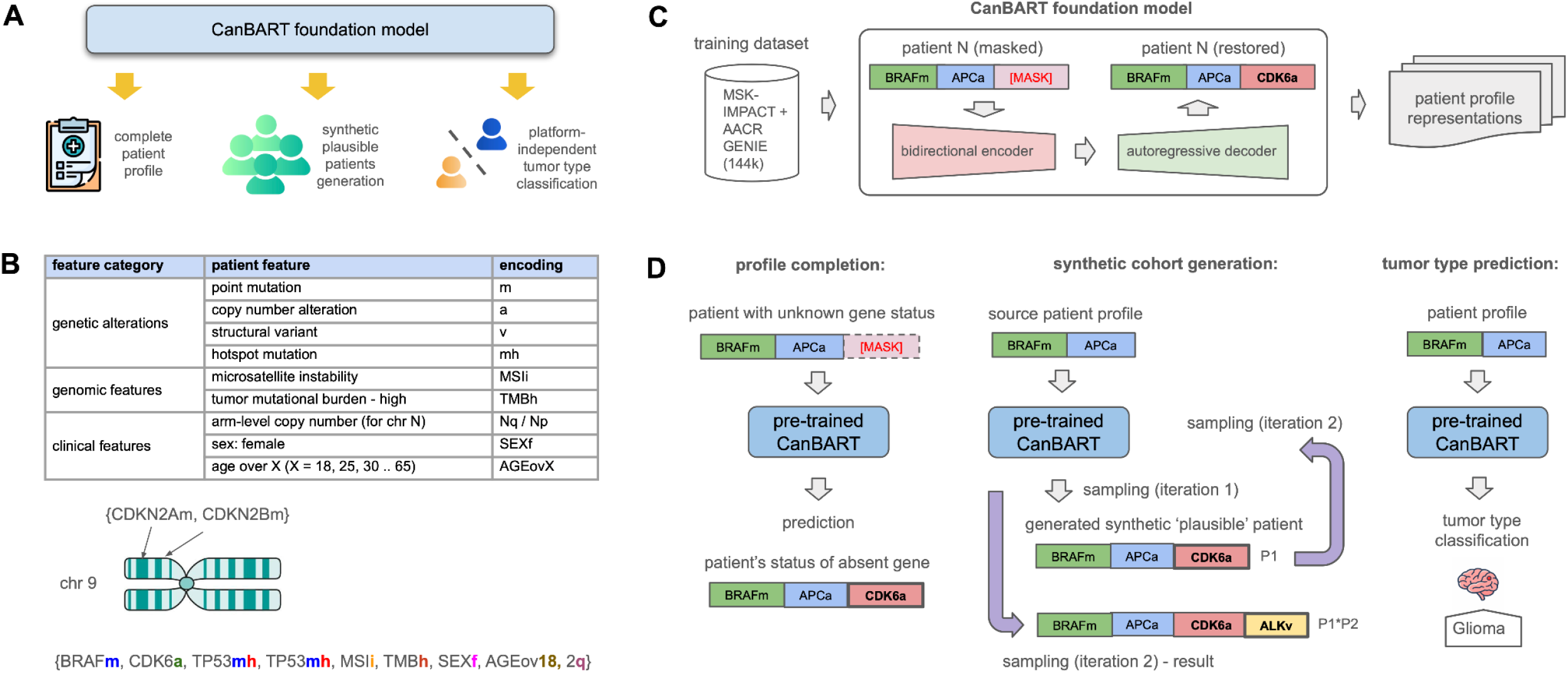
CanBART model overview, token construction, training architecture, and downstream use cases. **A)** Overview of downstream tasks supported by the CanBART foundation model. A single pre-trained model operates on patient-level clinico-genomic profiles and supports three core tasks: completion of incomplete genomic profiles via inference of missing gene-level alterations, generation of “plausible” synthetic patient profiles through probabilistic sampling from the learned alteration distribution, and platform-independent tumor type classification directly from genomic profiles, independent of the specific targeted sequencing panel. **B)** Input representation and patient profile encoding. Top: table summarizing gene-level molecular alterations and patient-level clinical attributes used as model inputs. Middle: illustration showing that alteration tokens are ordered according to the chromosomal positions of the corresponding genes. Bottom: example of a fully encoded patient profile, where each gene token is annotated with a suffix indicating the alteration type, and patient-level variables (e.g., MSI status, TMB category, sex, age group, and arm-level copy-number alterations such as 2q) are included as dedicated tokens. **C)** Training architecture of CanBART. The model is trained using a sequence-to-sequence BART architecture, in which partially masked patient profiles are processed by a bidirectional encoder and reconstructed by an autoregressive decoder. Masked-prediction objectives encourage the model to learn co-occurrence structure among molecular alterations and clinical features, resulting in biologically meaningful patient profile representations. **D)** Illustration of downstream use cases enabled by the learned patient representations. Left: Genomic profile completion, where a patient profile with missing gene status is provided as input and the model predicts the most likely alteration state for the missing gene. Middle: Synthetic patient generation, where the model iteratively samples additional alterations conditioned on the original patient profile. At each iteration, newly sampled alterations are added to the profile and the cumulative likelihood of the synthetic patient is computed as the product of stepwise probabilities. Right: Tumor type classification, where a complete or partial patient profile is mapped to a predicted tumor type label based on the learned representation.

To evaluate the model’s capabilities, we organized the analysis around the key tasks it supports in practice. We first assessed the masked-language modeling component in an imputation setting, testing whether the model could recover missing gene statuses in incomplete profiles and thereby simulate reduced targeted panels. We then evaluated whether the learned representations support tumor classification by distinguishing nine common cancer types from a pooled category of rare cancers, as well as by assessing separation among rare cancers in latent space. Finally, we examined the generative mechanism by producing “plausible” synthetic patients for rare cancer types via masked sampling and using these cohorts to augment training data, resulting in improved classification performance for rare cancers in low-data regimes. In summary, CanBART is a generative foundation model operating on patient-level clinico-genomic profiles that learns biologically meaningful representations of cancer and generates realistic synthetic cohorts, enabling cross-platform data harmonization across genomic profiling assays and improving tumor classification, particularly in underrepresented and data-limited settings.

## Methods

### Data Representation

Our study was based on a dataset primarily derived from the MSK-IMPACT panel, comprising genomic profiling results from approximately 80,000 patients (6). These profiles included three main categories of alterations: mutations, copy number alterations (CNA), and structural variants (SV), assessed across up to 505 cancer-related genes. Additional clinical attributes such as patient sex, age, microsatellite instability (MSI), and tumor mutational burden (TMB) were also available for subsets of the cohort. To broaden the dataset, we incorporated samples from the AACR GENIE database, which included genomic profiles for an additional 64,000 patients (7). To standardize representation across platforms, all profiles were restricted to the 505 genes covered by the full MSK-IMPACT panel; genes outside this maximal panel size were excluded.

To adapt Transformer architectures for clinical genomic data, we designed a sentence-like representation of the molecular alterations observed in patient tumor profiles. Instead of encoding profiles as binary feature vectors, which cannot capture multiple alterations affecting the same gene, we represented each profile as a sequence of alteration tokens sorted by chromosomal position. Each token contained the gene name and a suffix indicating the alteration type: mutation, hotspot, copy-number event, or structural variant (Fig. 1B). Alteration tokens are sorted by genomic position, so their order acts as a positional encoding: the location of each token in the sequence corresponds to its chromosomal coordinate. This ordering constrains the prediction space during masked-token inference: when a position is masked, the model selects from the limited set of alteration tokens that are valid for that genomic region (i.e., its restricted target list) rather than from the full vocabulary. In contrast, autoregressive sampling generates new tokens, but the same positional constraints ensure that only genes appropriate for that region can appear, keeping synthetic profiles biologically coherent.

### Model Architecture

We based our approach on BART, a sequence-to-sequence Transformer architecture with a bidirectional encoder and autoregressive decoder (Fig. 1C) (8). The model consisted of 3 encoder and 3 decoder layers, each with 1024-dimensional hidden states and 8 attention heads. A custom WordLevel tokenizer was trained on alteration tokens, resulting in a vocabulary of approximately 2,100 tokens.

The model was initially pre-trained using a masked language modeling (MLM) objective on over 144,000 tumor profiles from the MSK-IMPACT and AACR GENIE databases (Fig 1C). The MLM objective was designed to enable the model to learn combinatorial genomic patterns by reconstructing masked alterations in patient profiles represented as token sequences, and to build patient-level embeddings that support multiple downstream applications, similar to how foundation models are structured in NLP.

After pretraining, the model was fine-tuned for a tumor-type classification task (Fig. 2A). Patient profiles were mapped to 56 tumor types, each represented by at least 15 samples. Among these, nine cancer types with at least 6,000 samples each were designated as common, while the remaining 47 were considered rare and were analyzed in two separate experiments: first, as a single pooled rare cancer category contrasted against common cancers, in order to evaluate the model’s ability to distinguish rare from common types and to explore the extent of shared genomic signals, and second, as 47 distinct rare cancer classes to assess fine-grained classification performance.

**Fig. 2.**
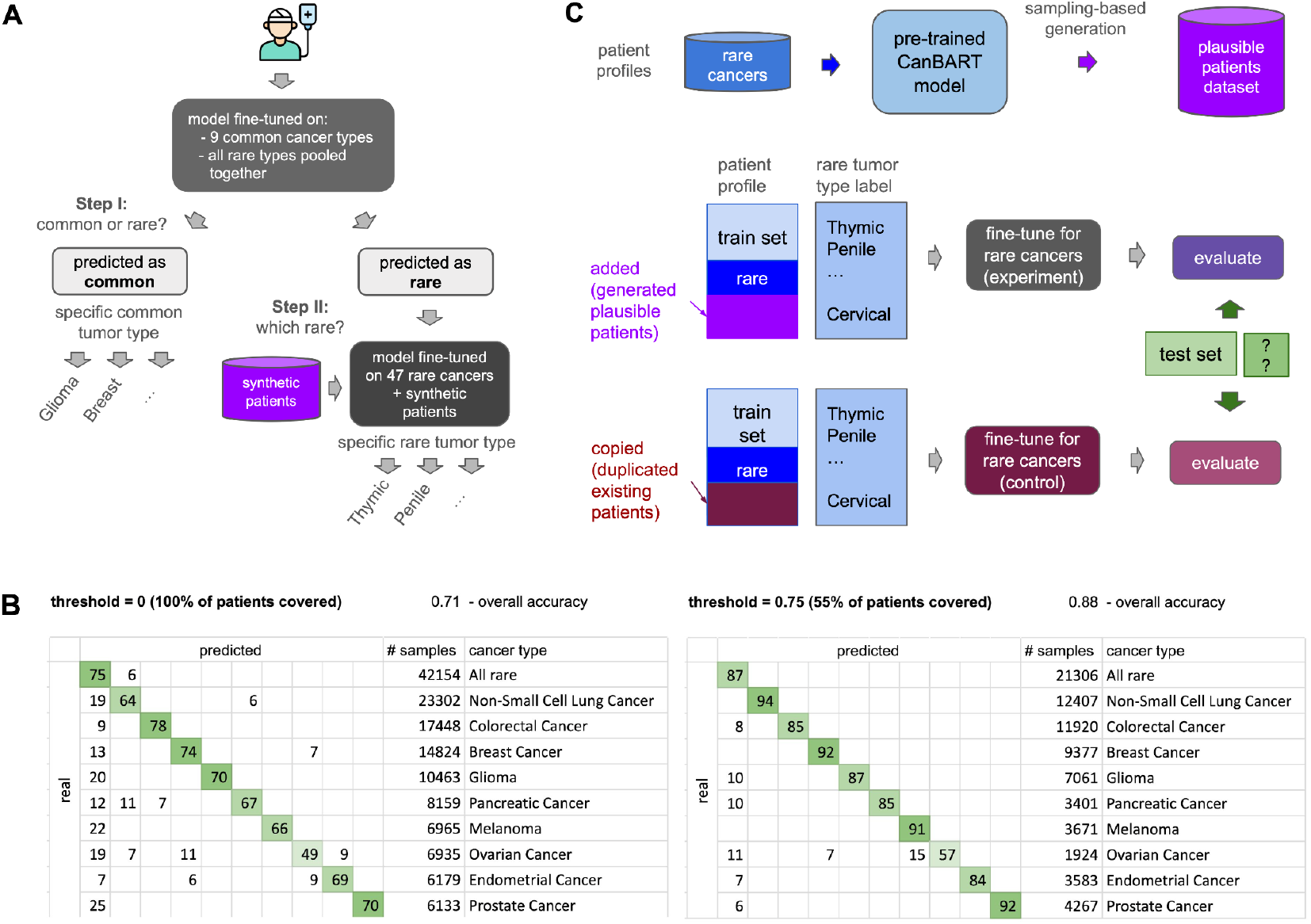
Two-stage tumor classification and synthetic data augmentation strategy for rare cancers. **A)** Two-stage tumor classification strategy based on CanBART. For tumor type classification, a patient clinico-genomic profile is first provided to a Stage I classifier fine-tuned on the 9 most common cancer types and a pooled rare-cancer category. Profiles classified as common are assigned a specific common tumor type. Profiles classified as rare are subsequently routed to Stage II, where a second classifier fine-tuned on 47 rare cancer types performs fine-grained rare tumor type classification. The Stage II classifier is trained with augmentation using synthetic patient profiles. **B)** Performance of Stage I common-versus-rare tumor classification. Confusion matrices are shown for classification of the 9 most common cancer types versus the pooled rare cancer group. Left: predictions for the full held-out test set. Right: predictions restricted to high-confidence cases (model probability > 0.75). Values ≤ 5 are omitted for clarity. **C)** Synthetic data augmentation strategy for rare cancer classification. Top: generation of plausible synthetic patient profiles for rare cancers using the pre-trained CanBART model, starting from real rare-cancer genomic profiles. Bottom: comparison of 2 fine-tuning strategies for the rare tumor classifier. In the augmentation branch, synthetic patient profiles are added to the rare-cancer training set. In the control branch, the original rare-cancer profiles are duplicated to match the size of the augmented dataset. Both classifiers are evaluated on the same held-out rare-cancer test set to enable a fair comparison of performance.

### Missing Gene and CNA Imputation

To assess the model’s capacity to infer unobserved genomic information, we simulated incomplete panel testing using the 278 genes shared with the DFCI panel. Profiles were restricted to these genes, and the model was evaluated on its ability to reconstruct the remaining 1,067 gene-level alterations across 181 genes from the MSK-IMPACT dataset. The pre-trained model was evaluated in a five-fold cross-validation setup, where in each fold the held-out test set was treated as a simulated DFCI panel and excluded from training. Missing genes were predicted in a binary classification setup, using the masked-token prediction probability as a confidence score. Performance was assessed using ROC-AUC across genes.

Beyond gene-level imputation, two additional tasks were evaluated within the same DFCI-panel restriction: (i) predicting arm-level CNAs (chromosomal gains and losses) from gene-level alteration patterns; and (ii) predicting gene-level CNAs from known arm-level CNA events. Both tasks used the same masked-prediction framework and were evaluated using ROC-AUC.

Together, these imputation tasks simulate real-world scenarios in which genomic panels provide only partial information. The model’s ability to infer missing alterations and CNA patterns supports clinical use cases such as therapy selection, genomic harmonization across panels, and trial matching.

### Plausible Patient Generation

To improve classification performance for rare cancers, especially those with limited data and insufficient training signal, we first sought to generate synthetic training examples for underrepresented tumor types (Fig. 1D). For this, we used the pre-trained model as a foundation, applying it without fine-tuning to generate new “plausible” profiles.

We implemented a masked autoregressive sampling strategy, inspired by text generation in NLP. Starting with a real patient profile, we iteratively masked one token at a time and sampled replacements from the model’s output distribution. Sampling followed a nucleus (top-p) strategy to promote diversity and avoid mode collapse.

Each generated profile was scored by its cumulative generation probability, calculated as the product of token probabilities at each generation step, where the model predicts the likelihood of each token at the masked position. A maximum of 50 iterative steps was used for generating synthetic patient profiles. Generation was stopped once the cumulative probability fell below a predefined threshold. This threshold was selected empirically for each cancer type by evaluating classification accuracy across multiple values, as described in the next section.

### Rare Cancer Classification with Synthetic Patients

We evaluated whether augmenting training data with synthetic rare cancer patients could improve classification performance (Fig. 2C). Rare cancers were defined as types with fewer than 6,000 samples and grouped into multiple size-based bins (e.g., 15-50, 50-150, 150-250, etc.). For each bin, experiments were conducted across varying numbers of cumulative probability thresholds, and, consequently, of the number of generated patients. The goal was to identify an optimal plausibility cutoff for each cancer type, based on classification accuracy.

Fine-tuning was performed using five-fold cross-validation. In each fold, a separate test set of real patient profiles was held out and not seen during model training. Accuracy was used as the evaluation metric and was reported per fold and averaged across all folds. The pre-trained model used as the base was trained on the full dataset prior to folding.

To ensure a fair comparison, we also tested a control condition where synthetic samples were replaced by duplicated real patients to keep dataset sizes equal.

## Results

### Missing Gene and CNA Imputation

We evaluated CanBART’s ability to impute missing genomic information in profiles restricted to the DFCI panel. For gene-level imputation, the model was tested on 1,067 masked alterations from MSK-IMPACT. Among alterations observed in at least 15 patients, 33% achieved ROC-AUC ≥ 0.75 (Fig. 3). Performance was lower for very infrequent events, reflecting their limited representation in the dataset.

**Figure 3.**
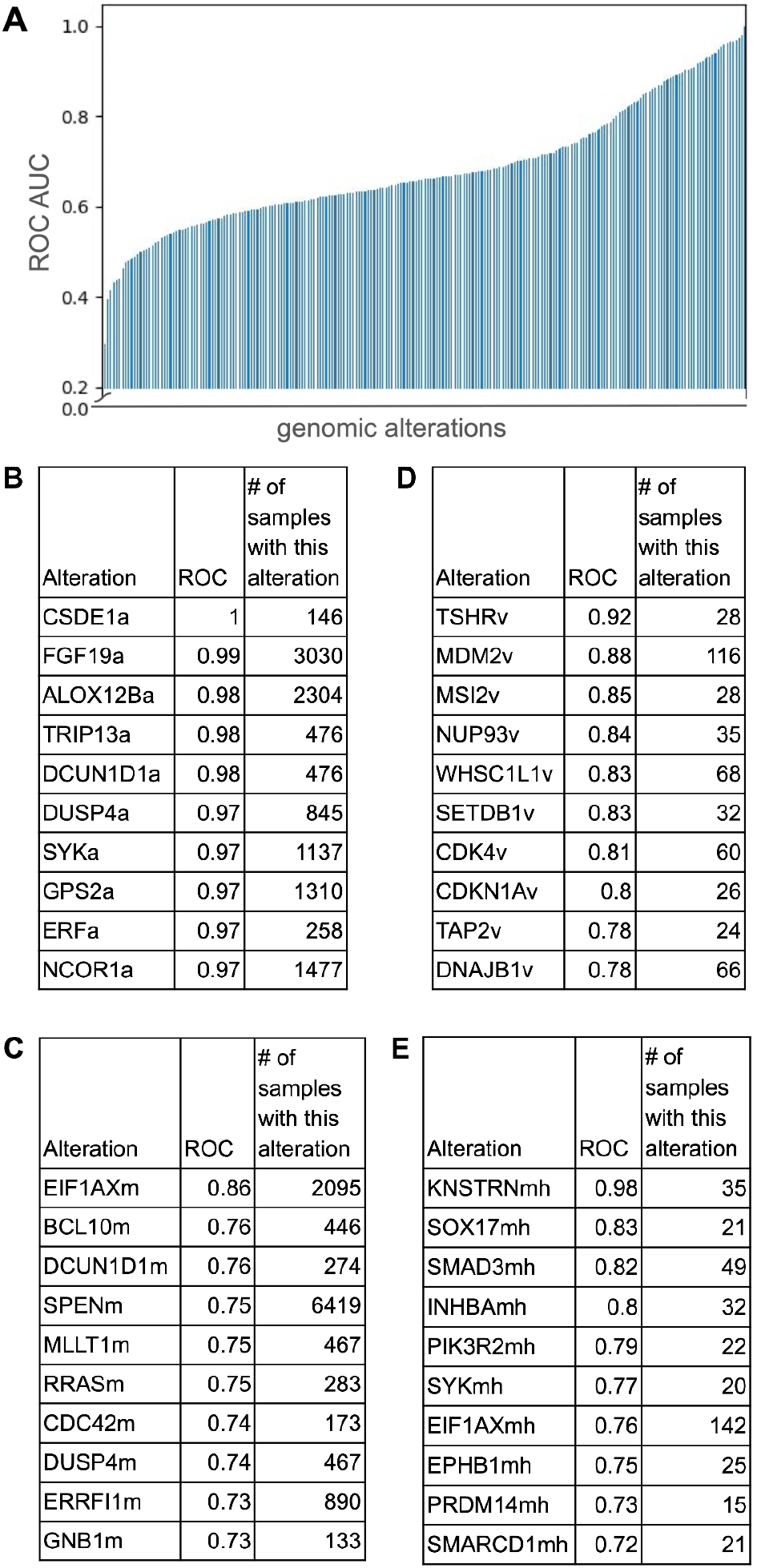
ROC AUC distribution and top-performing alterations in the gene status completion task. **A)** Histogram of ROC AUC scores for individual molecular alterations, sorted in ascending order. To simulate the DFCI targeted panel, genes not included in the panel were removed from each sample, and the model was trained to predict their status. Alterations with fewer than 15 positive training examples were excluded from evaluation, matching the threshold used in the classification task. Overall, 33% of alterations achieved ROC AUC ≥ 0.75. **B–E)** Top 10 most accurately predicted alterations, stratified by alteration type: **B)** copy-number alterations (CNAs); **C)** mutations; **D)** structural variants (SVs); **E)** hotspot mutations.

We next assessed whether the model could recover broader CNA patterns. Out of 46 arm-level CNA events, 21 (46%) were predicted with ROC-AUC ≥ 0.75. In the reverse task, predicting gene-level CNAs from known arm-level events, 41% of gene-level CNAs reached ROC-AUC ≥ 0.75. These results indicate that CanBART can reconstruct both fine-grained gene alterations and higher level chromosomal patterns from incomplete panels.

Together, these findings show that CanBART can harmonize partially sequenced profiles across multiple levels of genomic granularity and support downstream analyses in patients with incomplete or underprofiled sequencing data.

### Rare Cancer Classification

We evaluated classification performance in the pooled rare versus common cancer task. The 10-class confusion matrix is shown in Fig. 2B, and Fig. 2C displays the results for high-confidence predictions only (probability > 0.75). To visualize the internal structure of the learned representations, we projected the encoder’s hidden states using UMAP (Fig. 4) (9).

**Fig. 4.**
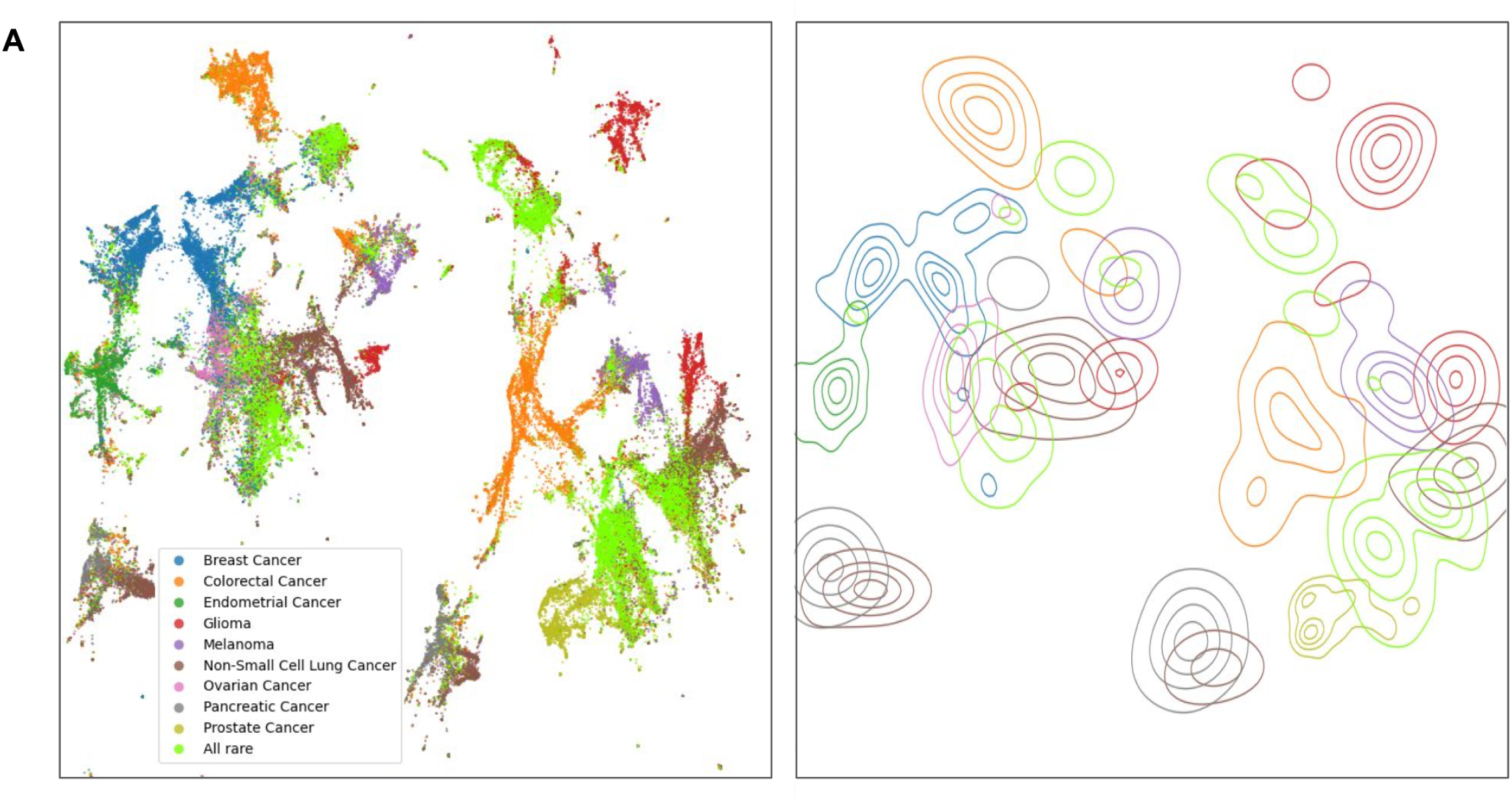
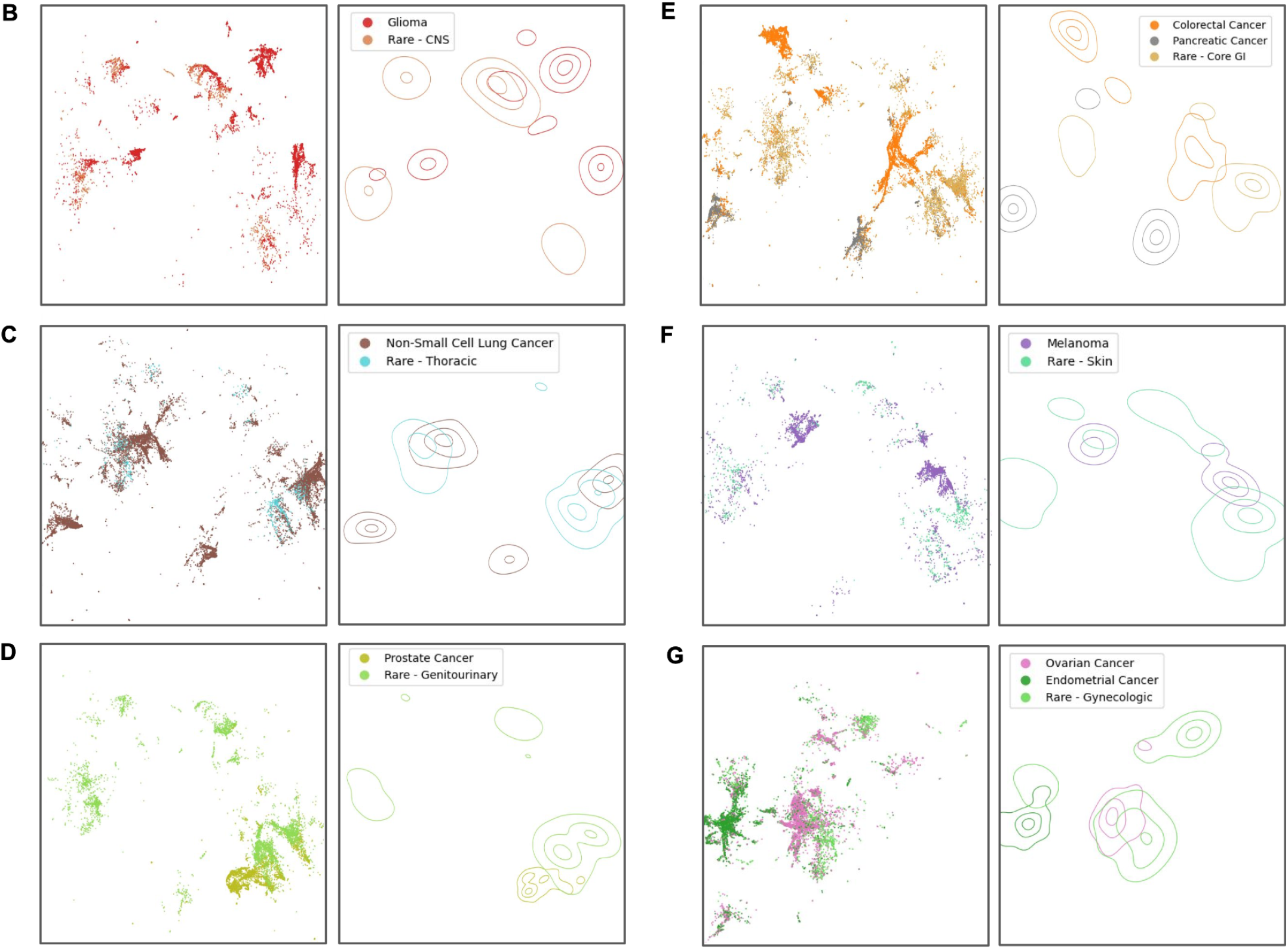
UMAP projections of CanBART patient representations for common and rare cancers. **A)** Global UMAP overview of patient representations. UMAP projection of latent patient representations for the nine most common cancer types and a pooled group of all rare cancers. Rare cancers are shown as a single group (left), highlighting their collective structure relative to common cancers. Right: corresponding density plots for the same cancer groups, illustrating the distribution of patient representations in the latent space. **B-G)** Organ-system–specific comparisons between common and rare cancers. For each panel, the left subpanel shows a UMAP projection and the right subpanel shows the corresponding density plot for common cancers and anatomically related rare cancers from the same organ system: **B)** gliomas and rare central nervous system (CNS) tumors; **C)** non–small cell lung cancer (NSCLC) and rare thoracic tumors; **D)** prostate cancer and rare genitourinary tumors; **E)** colorectal and pancreatic cancers with rare gastrointestinal tumors; **F)** melanoma and rare cutaneous tumors; **G)** ovarian and endometrial cancers with rare gynecologic tumors. Across organ systems, rare cancers occupy regions of the latent space that partially overlap with those of common cancers from the same anatomical context, indicating that CanBART captures shared molecular structure between common and rare tumor types

**Fig. 6.**
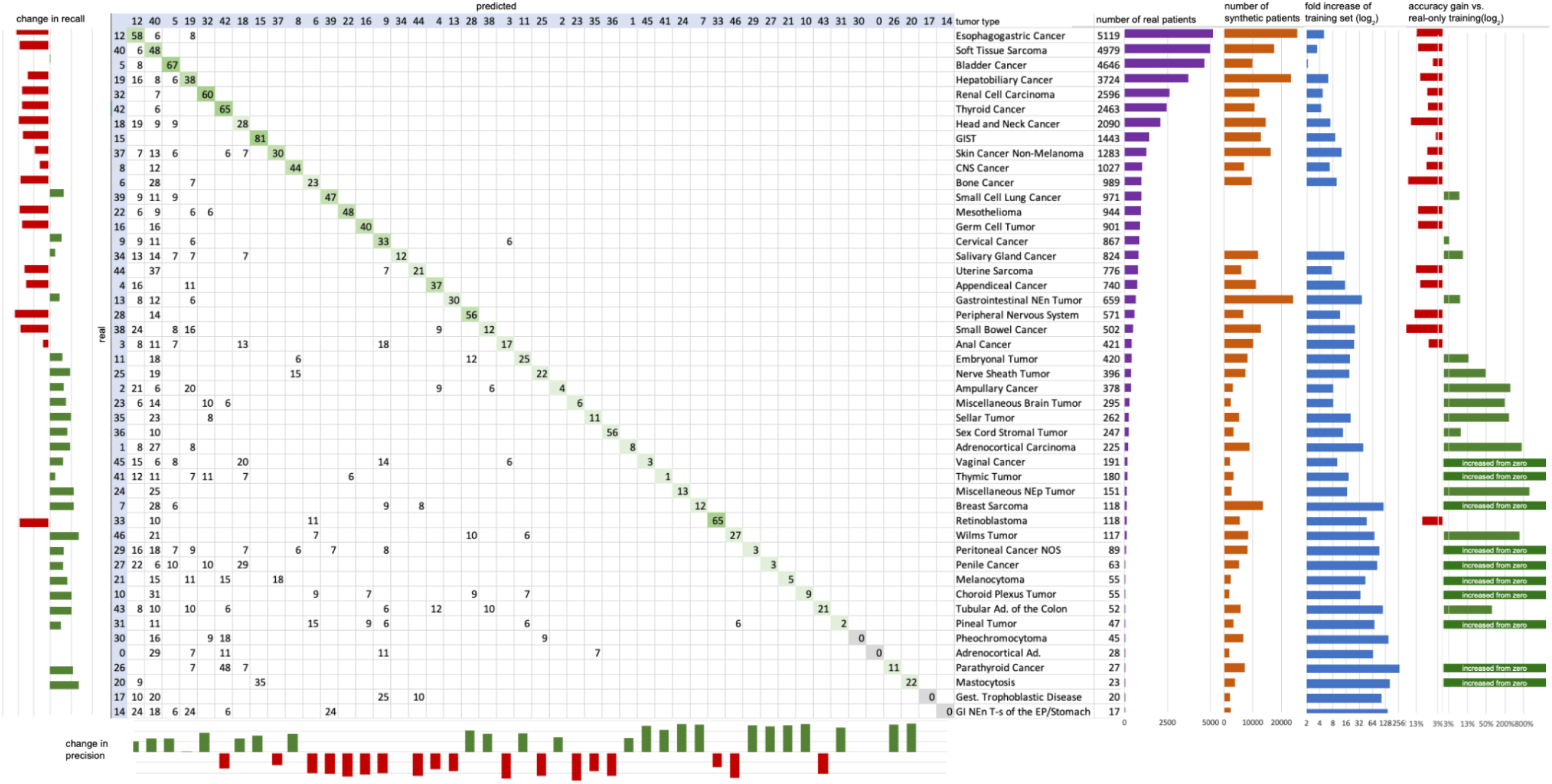
Performance of CanBART in rare cancer classification after synthetic patient augmentation. Confusion matrix showing classification performance across 47 rare cancer types following fine-tuning with synthetic patient augmentation. True cancer types are ordered from top to bottom by the number of available real patients per class. To the right of the confusion matrix, aligned summary panels provide additional context for each rare cancer type: **(i)** number of real (original) patients in the training set; **(ii)** number of synthetic patients generated for each cancer type using the optimization-guided augmentation strategy; **(iii)** fold increase in training set size after augmentation (log_2_ scale); **(iv)** changes in classification performance relative to the baseline model trained only on real patients, shown separately for precision and recall. Green indicates improvement and red indicates deterioration. This visualization illustrates that generative augmentation preferentially improves classification performance for ultra-rare cancers, many of which initially had very limited training data, while having limited or negative impact on better-represented rare cancer types.

This revealed that while some rare cancers cluster tightly, likely reflecting distinct molecular subtypes or well-represented rare types, many others are interspersed among common cancer clusters. These “signal-aligned” rare cancers appear adjacent to common types, suggesting latent proximity driven by shared genomic features and further supporting the idea that common cancers serve as anchors in the learned embedding space.

We then evaluated CanBART’s ability to classify 47 rare cancer types individually. The confusion matrix summarizing final performance after augmentation with synthetic patients is shown in Fig. 5. Per-class changes in classification accuracy are shown alongside the matrix, indicating that 28 cancer types exhibited improved accuracy, 14 showed decreased accuracy, and 5 remained unchanged. The number of synthetic patients added per cancer type (averaged across five folds) is also shown.

To control for dataset size, we conducted a parallel experiment using the same classification task across the 47 rare cancer types, where the number of training samples was augmented by simply duplicating existing real patients instead of generating new synthetic ones (Fig. 2C). Compared with the model trained using synthetic patient augmentation, classification performance under this duplication-based control was worse for 30 cancer types, with an average reduction in accuracy of 39% (95% CI: 27–50%). For 10 of the more prevalent rare cancer types, accuracy under duplication-based augmentation was slightly higher, though never by more than 6%, while for the remaining 7 types, performance was comparable between the two approaches.

Optimal cumulative probability thresholds used to select “plausible” synthetic patient numbers varied across cancer types and were determined empirically within bins defined by the number of available real patients.

## Discussion

This study presents CanBART, a Transformer-based foundation model trained on real-world genomic profiling data, designed to address key limitations associated with datasets for rare cancers and other underrepresented patient groups where available samples are insufficient for conventional modeling. By modeling molecular alterations as tokenized sequences and leveraging a BART-based architecture, CanBART bridges generative modeling in the clinical cancer genomics domain.

A core result of our study is that representations learned from common cancer types can generalize to rare ones. This was demonstrated in pooled classification and latent space visualization: many rare tumors clustered near common cancers, suggesting the presence of transferable genomic signals. These signal-aligned rare cancers may share driver events, mutational patterns, or structural variants with more prevalent tumor types, allowing the model to anchor their representations through shared latent features. This supports prior hypotheses proposed in the GDD-ENS study and offers quantitative evidence that Transformer-based models can exploit implicit similarity across cancer types for data-efficient learning (5).

Importantly, this shared signal is not only visible in the latent structure: it also manifests functionally. Synthetic patients generated by CanBART, inheriting patterns learned from common tumors, led to measurable accuracy improvements when added to training data for rare cancer classification. This demonstrates that the generated profiles carry meaningful, cancer-specific signals rather than generic noise. In our setup, the classification task was not intended to yield a clinically deployable classifier, but rather to validate the informativeness of synthetic examples. Performance for some rare types remained modest, largely due to extremely small training cohorts.

However, the consistent gains achieved through generative augmentation justify the use of “plausible” patients in tasks where annotated datasets are scarce. These synthetic cohorts could support a wide range of downstream applications in settings where real-world data is limited. Crucially, the success of this approach confirms that the sentence-like representation of patient data, composed of molecular alterations and key clinical attributes is expressive enough to capture both tumor identity and the molecular portrait of individual cases. This implies that synthetic patients can participate meaningfully in digital experimentation and serve as useful proxies in planning clinical trials or modeling therapeutic response. They may be used to discover novel biomarkers by revealing consistent alteration patterns within rare subtypes, to simulate therapy response distributions *in silico* for trial planning, or to create virtual control arms for clinical studies. Moreover, they could serve as training data for predictive models in settings like progression-free survival, resistance profiling, or drug repurposing for underrepresented cancer populations. As models like CanBART become more refined, synthetic cohorts may also facilitate functional hypothesis generation, safety profiling, and prioritization of targets for experimental validation.

The observed classification performance should be interpreted in the context of several compounding factors. First, the dataset used for training included not only MSK profiles but also samples from dozens of institutions via the AACR GENIE consortium (7). This increased heterogeneity makes the model more generalizable but also introduces substantial batch effects. Second, the platform diversity within AACR GENIE is substantial, with over a hundred gene panels represented, some as small as 50 genes. This variability likely contributes to misclassification, particularly among patient profiles with minimal profiling coverage. Importantly, the strongest accuracy improvements from synthetic patient augmentation were observed in cancers with 20 to 500 samples, indicating that the generative model is most effective in filling low-data regimes. However, we also observed some misclassification among more common cancers, likely due to signal entanglement introduced by augmentation. One potential solution may involve stratified training, with separate classifiers optimized for ultra-rare, rare, and common cancer groups to mitigate interference between signal-rich and signal-poor classes.

To further examine the biological grounding of our generative mechanism, we evaluated whether the same architecture could be used to predict unobserved molecular alterations. In a panel reduction setting, CanBART accurately recovered a substantial fraction of masked events, suggesting that the model internalized co-occurrence patterns from the training corpus. In addition to gene-level events, the same framework recovered a large subset of chromosome arm–level CNA events with high ROC-AUC and correctly inferred a substantial fraction of gene-level CNAs from arm-level signals, indicating that the model can reconstruct both local and higher-order copy-number patterns from incomplete panels. This not only supports the plausibility of the synthetic profiles used in classification tasks but also demonstrates the model’s utility for gene-level inference. This capability is becoming increasingly important given the diversity of sequencing platforms in clinical use and the fragmentation of genomic coverage across institutions. Foundation models like CanBART offer a promising route toward cross-platform harmonization, enabling robust comparisons and unified representation across incomplete profiles.

## Conclusion

Together, these results establish CanBART as a flexible foundation model for genomic oncology and computational drug discovery: capable of classifying rare tumors, simulating plausible cohorts, and imputing incomplete profiles using a shared generative engine. More broadly, our work illustrates how generative pretraining on real-world cancer data can support a spectrum of downstream applications without requiring handcrafted features or extensive per-task engineering. As genomic data collection continues to scale, foundation models like CanBART may become essential infrastructure for harmonized learning, interpretation, and synthetic data generation in precision oncology.

## Data Availability

All data produced in the present study are available upon reasonable request to the authors

## Acknowledgments

We thank Daniel Kelly and the Data & Analytics Group (DiGits, MSK) for their assistance in obtaining and managing the IMPACT molecular event dataset used in this study.

## References

1. Jumper, J., Evans, R., Pritzel, A. et al. Highly accurate protein structure prediction with AlphaFold. Nature 596, 583–589 (2021). 10.1038/s41586-021-03819-2.

2. Avsec, Ž., Agarwal, V., Visentin, D. et al. Effective gene expression prediction from sequence by integrating long-range interactions. Nat Methods 18, 1196–1203 (2021). 10.1038/s41592-021-01252-x.

3. Chen J, Hu Z, Sun S, Tan Q, Wang Y, Yu Q, et al. Interpretable RNA foundation model from unannotated data for highly accurate RNA structure and function predictions. arXiv. 2022. Available from: https://arxiv.org/abs/2204.00300.

4. Fradkin P, Shi R, Dalal T, Isaev K, Frey BJ, Lee LJ, Morris Q, Wang B. Orthrus: Towards Evolutionary and Functional RNA Foundation Models. bioRxiv [Preprint]. 2025 Jul 27:2024.10.10.617658. doi: 10.1101/2024.10.10.617658.

5. Darmofal M, Suman S, Atwal G, Toomey M, Chen JF, Chang JC, Vakiani E, Varghese AM, Balakrishnan Rema A, Syed A, Schultz N, Berger MF, Morris Q. Deep-Learning Model for Tumor-Type Prediction Using Targeted Clinical Genomic Sequencing Data. Cancer Discov. 2024 Jun 3;14(6):1064–1081. doi: 10.1158/2159-8290.CD-23-0996.

6. Zehir A, Benayed R, Shah RH, Syed A, Middha S, Kim HR, et al. Mutational landscape of metastatic cancer revealed from prospective clinical sequencing of 10,000 patients. Nat Med. 2017;23(6):703–713. doi:10.1038/nm.4333.

7. AACR Project GENIE Consortium. AACR Project GENIE: Powering Precision Medicine through an International Consortium. Cancer Discov. 2017;7(8):818–831. doi:10.1158/2159-8290.CD-17-0151.

8. Lewis M, Liu Y, Goyal N, Ghazvininejad M, Mohamed A, Levy O, et al. BART: Denoising Sequence-to-Sequence Pre-training for Natural Language Generation, Translation, and Comprehension. ACL. 2020;7871–7880. doi:10.18653/v1/2020.acl-main.703.

9. McInnes et al., (2018). UMAP: Uniform Manifold Approximation and Projection. Journal of Open Source Software, 3(29), 861, 10.21105/joss.00861.

